# From Advice to Action: Real-World Behaviour of Patients Using an Integrated Clinical AI for Navigating the Healthcare System

**DOI:** 10.1101/2025.07.24.25332138

**Authors:** Fabienne Cotte, Filipa Dias Lourenço, Miguel Paiva Pereira, Nisha Kini, Marcel Schmude, Andreia Pimenta, Athena Lemesiou, Stephen Gilbert, Tauseef Mehrali, Micaela Seemann Monteiro, Pedro Flores

## Abstract

**Background:** AI-powered digital front-door (DFD) tools are increasingly used to guide patients to appropriate care and alleviate healthcare system pressure. Yet, most evaluations narrowly assess triage accuracy or stated intent, offering limited insight into real-world behaviour and care appropriateness.

**Methods:** The ESSENCE study was a prospective, real-world quality improvement evaluation embedded in Portugal’s largest private healthcare network (CUF). Adults using the Ada Health clinical AI (cAI) via the myCUF app reported their care intent before and after symptom assessment. Actual behaviour was tracked through electronic health records and follow-up surveys. Physician panels retrospectively assessed the appropriateness of intended and observed care.

**Results:** Of 1,470 adult (≥18 years) participants (mean age 38.5 years; 57.7% female), 33% revised their planned choice of care immediately after assessment. Uncertainty dropped from 13% to 5%. Among 721 participants with follow-up, 59% changed their behaviour: 29% de-escalated, 17% escalated, and 13% resolved prior uncertainty. General practitioner (GP) visits more than doubled (42% vs. 19%), while specialist visits halved (30% vs. 57%). Overall, the proportion of appropriate care increased from 30% pre-assessment to 64% post-behaviour (p□<□0.001). Of those initially planning to visit the emergency department, 39% opted for alternative care. 93% of those followed up were later confirmed to have avoided unnecessary emergency department (ED) visits.

**Conclusion:** Embedding AI symptom assessment into a DFD shifted both patient intentions and behaviours, reducing uncertainty and promoting more appropriate healthcare use. These findings highlight the value of AI not only in generating accurate recommendations, but also in shaping real-world decision-making. Future research should identify specific factors that influence behaviour and evaluate impact across diverse healthcare settings.

## Introduction

Healthcare systems worldwide face growing strain from rising chronic disease burdens, ageing populations, and shortages of healthcare professionals. These pressures stretch resources, increase wait times, and limit access to timely care^1^. To address these challenges, scalable solutions are needed that guide patients to the right care at the right time - without compromising quality or safety.

One such solution is the *digital front door* (DFD): digitally enabled entry points through which patients access healthcare services. These tools - including symptom assessments, online triage, appointment booking, and remote consultations - are designed to streamline access, reduce unnecessary demand, and guide users to the most appropriate care setting^2^. Critically, they hold potential to support patients in one of the most challenging aspects of healthcare: deciding if, when, where, and how urgently to seek care.

Evidence shows that this decision-making process is filled with uncertainty. Self-triage accuracy remains moderate (47–62%), with frequent overestimation of urgency and occasional failure to recognise serious conditions^3,4^. While most individuals correctly identify emergencies (82%), overtriage occurs in 65% of cases, and 8% of critical situations are missed entirely^5^. As highlighted during the COVID-19 pandemic, delayed or misdirected care can lead to worse health outcomes^6^.

In response, healthcare systems are increasingly embedding AI-based symptom assessment and navigation into patient-facing platforms. These integrations aim to reduce uncertainty, curb unnecessary demand, and improve timely access to care.

Yet, most evaluations of digital triage tools focus narrowly on output accuracy or stated intent, offering limited insight into actual patient behaviour. For example, a study in Portugal reported that 22.8% of users revised their care plans after AI-supported triage, but did not assess whether patients followed through with their intention or whether the resulting care was appropriate^7^. Similarly, an evaluation in Australia described reductions in overtriage compared to the previous system but did not examine adherence to recommendations or downstream outcomes^8^. These approaches miss the complex, often non-linear ways patients interpret information and make decisions in real-world settings.

In reality, users are often exposed to more than a single triage recommendation. They receive a mix of condition-specific advice, explanatory information, and follow-up options - all of which, alongside personal and contextual factors, influence their decisions. Understanding this interplay requires evaluations that go beyond intent shifts to track actual behaviour and clinical appropriateness.

To address this gap, we conducted the ESSENCE study (E-health Self Symptom assEssmeNt as a front door and facilitator of CarE). Embedded within Portugal’s CUF private healthcare network, the study assessed how integrating Ada Health’s clinical AI into a DFD influenced patient behaviour, service use, and care appropriateness in a real-world setting.

### Objectives

This paper focuses on the behavioural impact of the Ada-CUF integration. Specifically, we assess how AI-driven symptom assessment affects patients’ healthcare-seeking decisions, actual service utilisation, and the appropriateness of resulting care. Findings related to clinical workflow and physician perspectives will be reported separately.

## Methods

### Study Design and Setting

This prospective, observational quality improvement study evaluated the real-world impact of a clinical AI (cAI, Ada) on healthcare-seeking behaviour within Portugal’s largest private healthcare network (CUF). Adults (≥18 years) using the myCUF app in Portuguese to complete a full symptom assessment for themselves were eligible. Inclusion required completion of pre- and post-assessment questions, informed consent, and report sharing with a healthcare provider. Participants were instructed not to use the cAI in emergency situations. Enrolment ran from November 2023 to October 2024.

### Data Collection Procedures

Participants reported their care-seeking intention before starting the assessment and again after reviewing the symptom report, which listed likely conditions with probability ratings and care recommendations for each condition and the overall case. Each condition included a “What’s next?” button linking to follow-up service options (e.g. booking a teleconsultation), though these were only shown after the post-assessment intention question was completed.

Healthcare-seeking behaviour was determined via electronic health record (EHR) review for consultations within CUF up to 14 days post-assessment. For those without a documented consultation, an email follow-up was sent 2–4 weeks later to collect self-reported care use outside CUF or home management with the same care options as listed in the intent survey.

For CUF consultations, additional data were extracted (clinical notes, ICD-10 codes, treatment etc.). If the participant was seen by a physician participating in the study at one of three participating CUF centres, a survey evaluating the symptom report was completed directly after the consultation or, if necessary, at a later time point.

At study end, a panel of three senior CUF physicians retrospectively reviewed each healthcare visit to determine the most appropriate level of care (self-care, primary care, specialist care, or emergency care).

ED visit appropriateness was assessed separately from non-ED visits using a three-part framework (Naouri et al., 2020), evaluating clinical appropriateness, potential for primary care evaluation, and type of resource use in the ED. Visits were classified as appropriate if they were either clinically justified or required ED-specific interventions.

### Data Analysis

All analyses were conducted in Python (v3.12). Descriptive statistics summarised participant demographics, care intentions, observed behaviours, and cAI recommendations. Continuous variables were reported as means (±SD) or medians (IQR); categorical variables as frequencies and percentages.

Care transitions across timepoints (pre-assessment, post-assessment, observed behaviour) were visualised with Sankey diagrams. Agreement between intended and actual behaviour was assessed using Cohen’s □; changes in care distributions using the Stuart–Maxwell test. McNemar’s test evaluated shifts in care selections across timepoints, and associations between cAI advice and behaviour were tested using Pearson’s χ^2^.

System-level utilisation impact was assessed by assigning ordinal weights to care levels (Self-care = 0, GP = 1, Specialist = 2, Emergency care = 4). Mean intended and actual resource intensity scores were then compared using the Wilcoxon signed-rank test. Differences between adherent (intention = behaviour) and non-adherent users were analysed using the Mann–Whitney U test.

Univariate analyses explored predictors of plan change, using χ^2^ for categorical and Mann– Whitney U for continuous variables. Appropriateness of intentions and behaviours was benchmarked and analysed with McNemar’s exact test.

### Generalisability

To assess selection effects, we compared care intentions and user characteristics between the study population and all other cAI users who completed an assessment within the same integration (excluding skipped responses). Within the study population, we examined differences between participants with and without follow-up data, including demographics, cAI-recommended care level, and changes in intended care.

### Data Management and Ethics

Data was securely stored in a validated electronic system (Teamscope) using unique identifiers. Ethical approval was granted by the Comissão de Ética para a Investigação Clínica (No. **2204JJ351** and No. **2309JJ660**). The study was registered at ClinicalTrials.gov (NCT06846957) and complied with the Declaration of Helsinki and ISO 14155:2020 guidelines. STROBE guidelines were followed throughout.

## Results

### Participant Demographics

The study enrolled 1470 participants, with a mean age of 38.5 years (SD 12.5); 57.7% (848/1470) were female. Participants initially entered a median of 2 symptoms (IQR 1–3; mean 2.3 ± 1.9). The cAI advised primary care in 74.8% (1,099/1,470) of cases. Healthcare visit data were available for 49.1% of participants (721/1,470). Among those who sought care (671/721), 31.4% (211/671) used teleconsultations (Table 1). The participant selection and inclusion process is outlined in a STROBE flowchart (Figure 1).

**Table 1.**
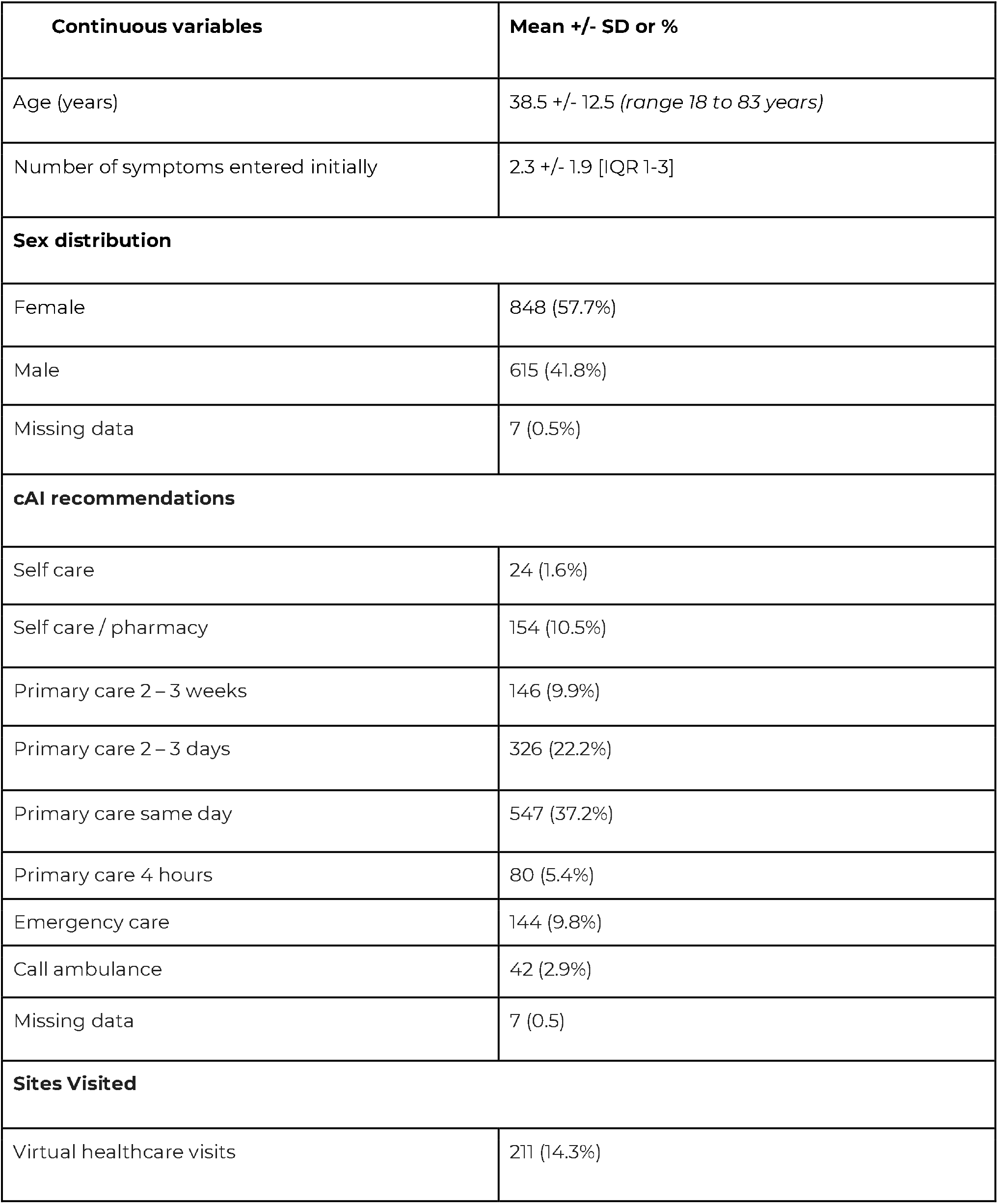
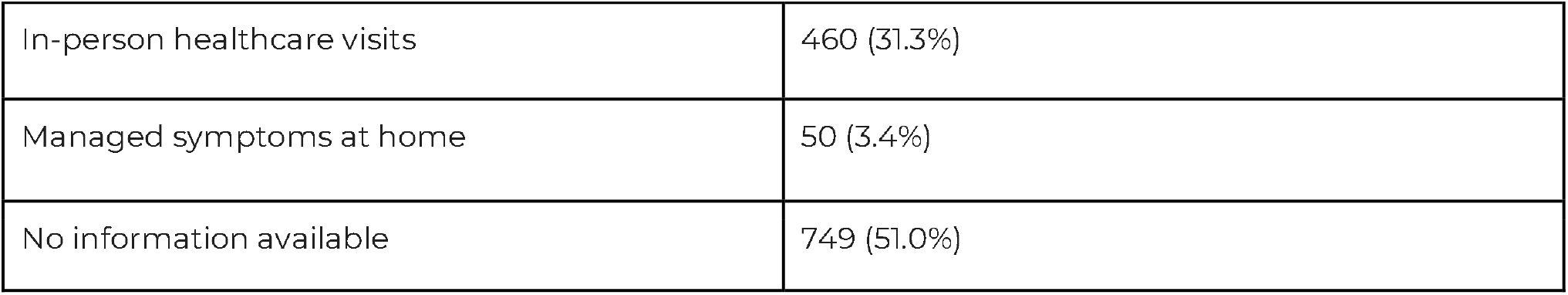
Participant Demographics (N = 1,470)

| Continuous variables | Mean +/- SD or % |
| --- | --- |
| Age (years) | 38.5 +/- 12.5 (*range 18 to 83 years*) |
| Number of symptoms entered initially | 2.3 +/- 1.9 [IQR 1-3] |
| **Sex distribution** |  |
| Female | 848 (57.7%) |
| Male | 615 (41.8%) |
| Missing data | 7 (0.5%) |
| **cAI recommendations** |  |
| Self care | 24 (1.6%) |
| Self care / pharmacy | 154 (10.5%) |
| Primary care 2 – 3 weeks | 146 (9.9%) |
| Primary care 2 – 3 days | 326 (22.2%) |
| Primary care same day | 547 (37.2%) |
| Primary care 4 hours | 80 (5.4%) |
| Emergency care | 144 (9.8%) |
| Call ambulance | 42 (2.9%) |
| Missing data | 7 (0.5) |
| **Sites Visited** |  |
| Virtual healthcare visits | 211 (14.3%) |

| **Continuous variables** | **Mean +/- SD or %** |
| --- | --- |
| In-person healthcare visits | 460 (31.3%) |
| Managed symptoms at home | 50 (3.4%) |
| No information available | 749 (51.0%) |

**Figure 1:**
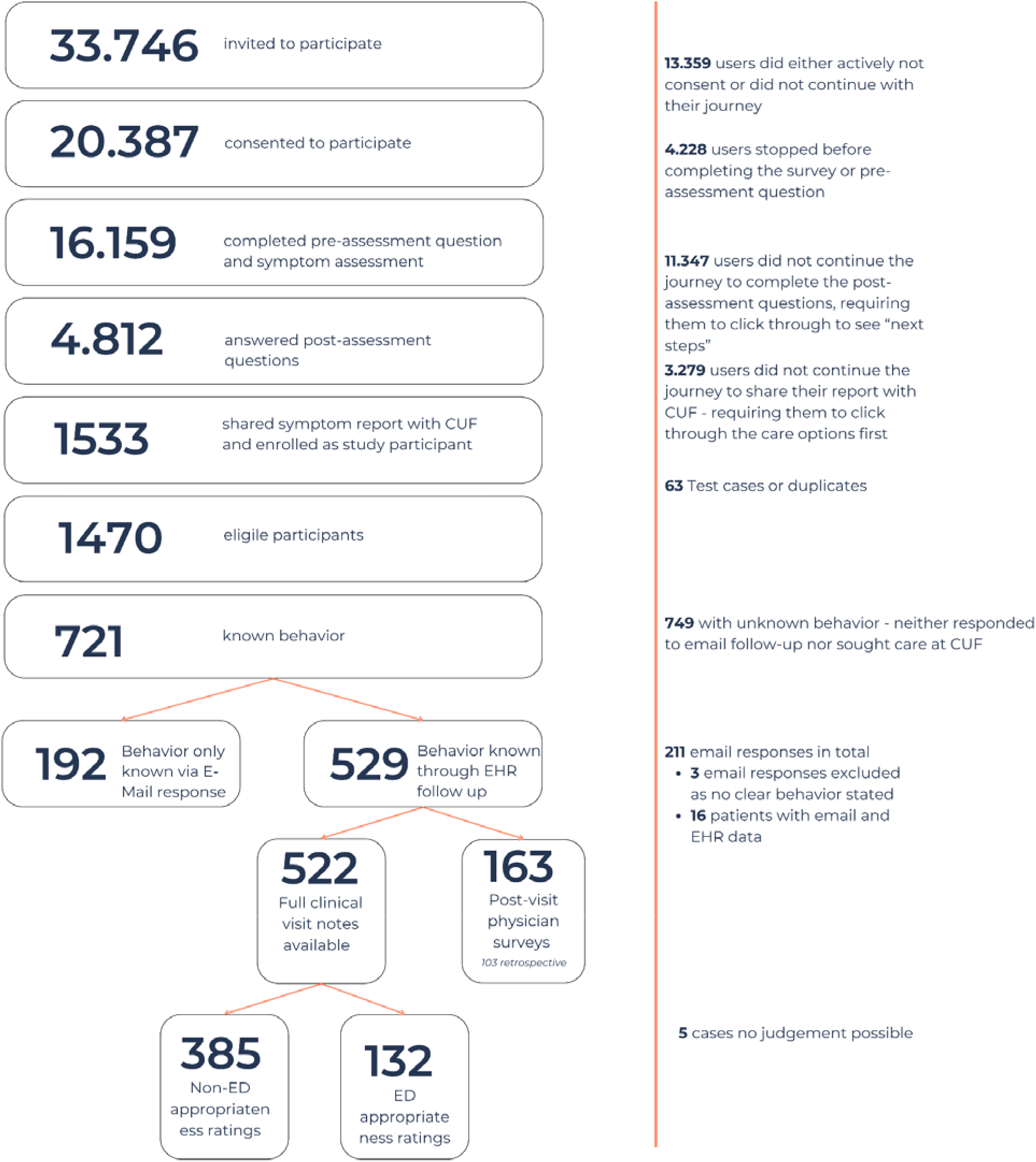
STROBE Inclusion flowchart

### Participant Intentions (Pre-vs Post-Assessment)

Participant intentions are summarised in Tables 2 and 3 and visualised in Figure 2A. Post-assessment, uncertainty declined significantly to 5.0□% (68/1351; p□< □0.001), a 61.5□% relative reduction. Intentions to consult a specialist increased from 52.9□% to 57.4□% (p□< □0.001), and to see a GP from 16.9□% to 19.5□% (p□= □0.082). The proportions planning to seek emergency care (10.7□%; p□=D0.053) or manage symptoms independently (7.3□%; p□= □1.000) remained largely unchanged. The overall distribution of urgency categories shifted significantly (p□< □0.001), with only 67.1□% (898/1338) of participants maintaining their original intention.

**Table 2.**
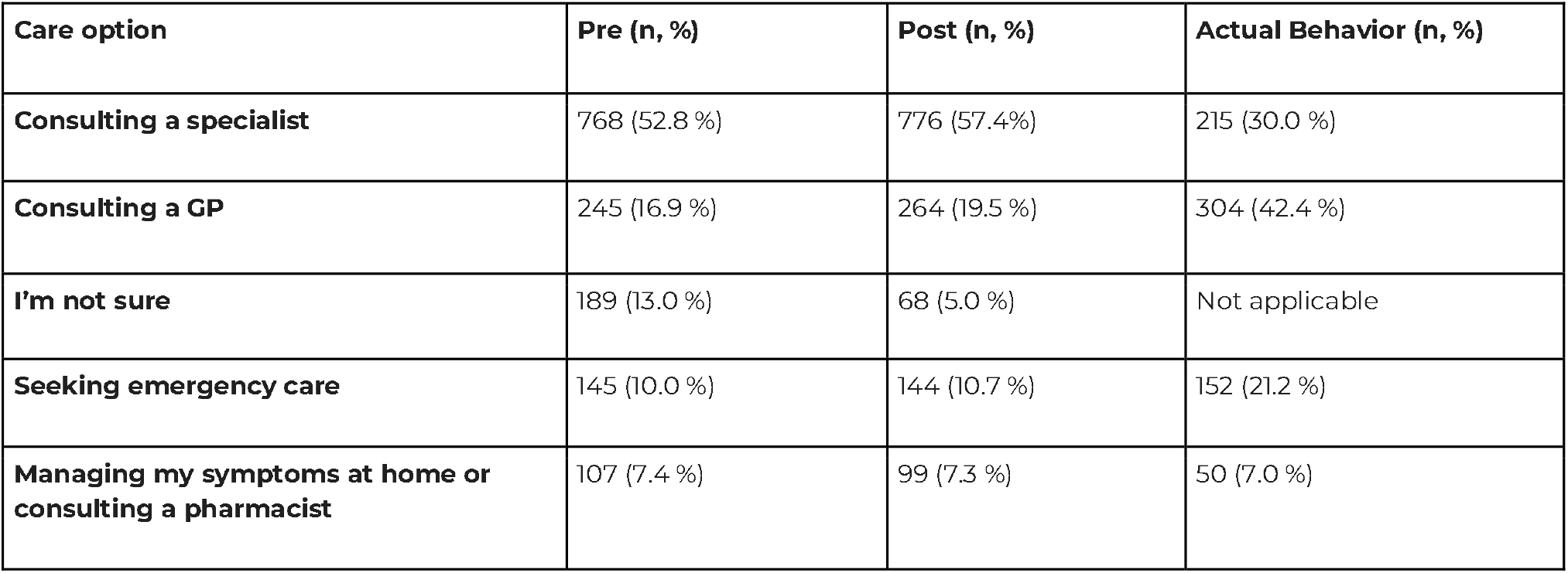
Care Level chosen before and after the symptom assessment and actual behavior, Pre-assessment total n = 1454; Post-assessment total n = 1351; Actual Behavior total n = 721. Excluding skipped responses or missing data for user behavior.

**Table 3.** Transition outcomes for pre-vs post-assessment intentions and observed behaviour.

| Metric | Pre → Post (n = 1338) | Pre → Behavior (n = 717) | Post → Behavior (n = 659) |
| --- | --- | --- | --- |
| **No change** | 897 (67.0 %) | 294 (41.0%) | 299 (45.4%) |
| **Escalated\*** | 126 (9.4 %) | 123 (17.2 %) | 122 (18.5%) |
| **De-escalated\*** | 110 (8.2 %) | 207 (28.9%) | 209 (31.7 %) |
| **Made certain†** | 138 (10.3 %) | 93 (13.0 %) | 29 (4.4 %) |
| **Made uncertain†** | 37 (2.8 %) | 0 (0.0 %) | 0 (0.0 %) |
| **Stayed uncertain‡** | 30 (2.2 %) | 0 (0.0 %) | 0 (0.0 %) |
| **Stuart-Maxwell test of marginal homogeneity** | $Q = 48.20$, $df = 4$, $p < 0.001$ | $Q = 124.4$, $df = 6$, $p < 0.001$ | $Q = 90.39$, $df = 3$, $p < 0.001$ |
| **Cohen's Kappa** | Cohen's $\kappa = 0.52$ | Cohen's $\kappa = 0.25$ | Cohen's $\kappa = 0.25$ |

\*Escalated = moved to a higher-urgency category; De-escalated = moved to a lower-urgency category.
†Made certain = "I'm not sure" → specific category; Made uncertain = specific category → "I'm not sure."
‡Stayed uncertain = "I'm not sure" at both timepoints,

**Figure 2A:**
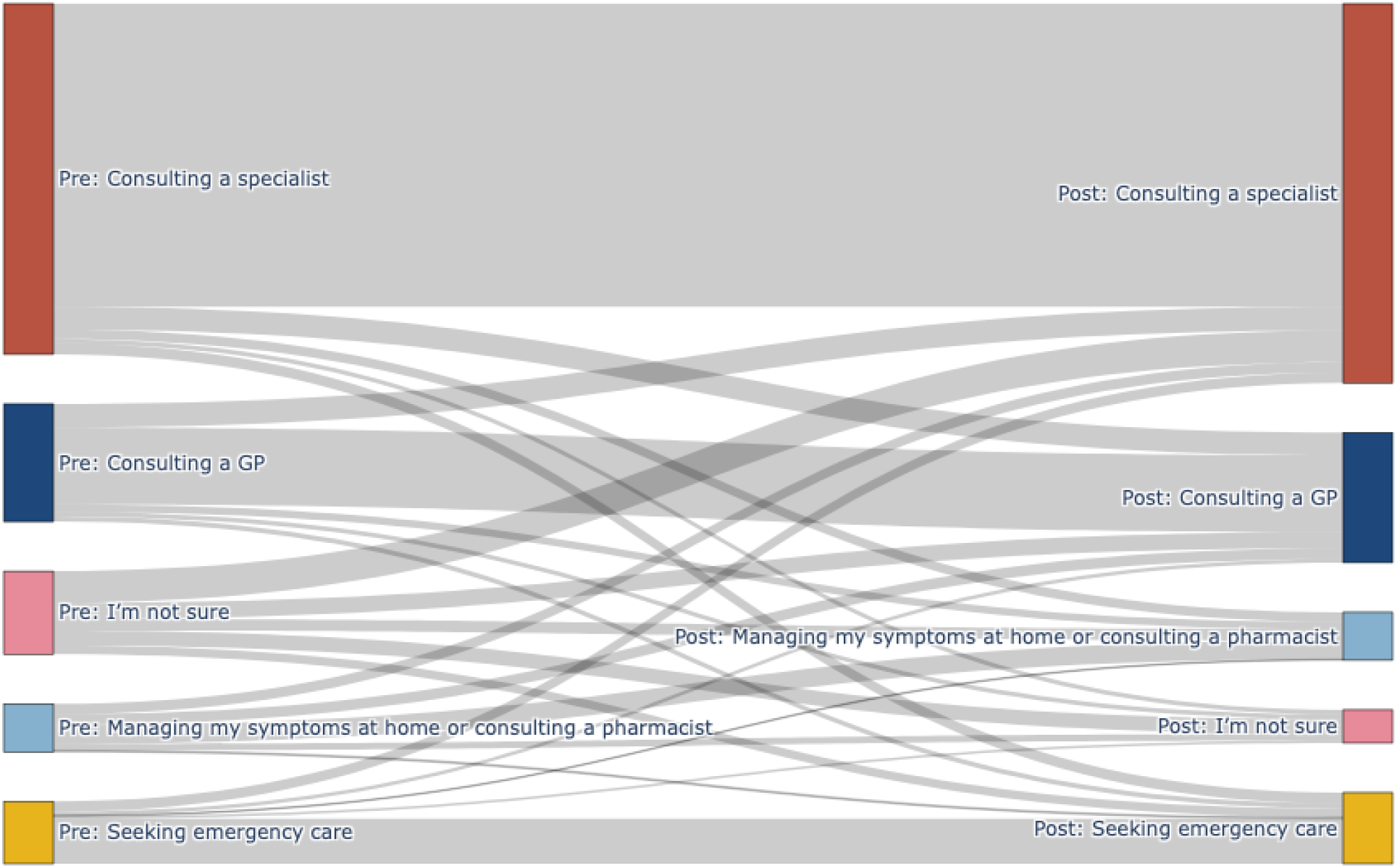
Sankey diagram depicting transitions between merged care categories from pre-to post-assessment intention. Node colors correspond to care categories: Consulting a GP (blue), Consulting a specialist (red), Managing home care/see pharmacist (light blue), I’m not sure (pink), Seeking emergency care (yellow). Link widths are proportional to the number of participants moving between categories. Participants with “Skip” responses at either timepoint were excluded.

**Figure 2B:**
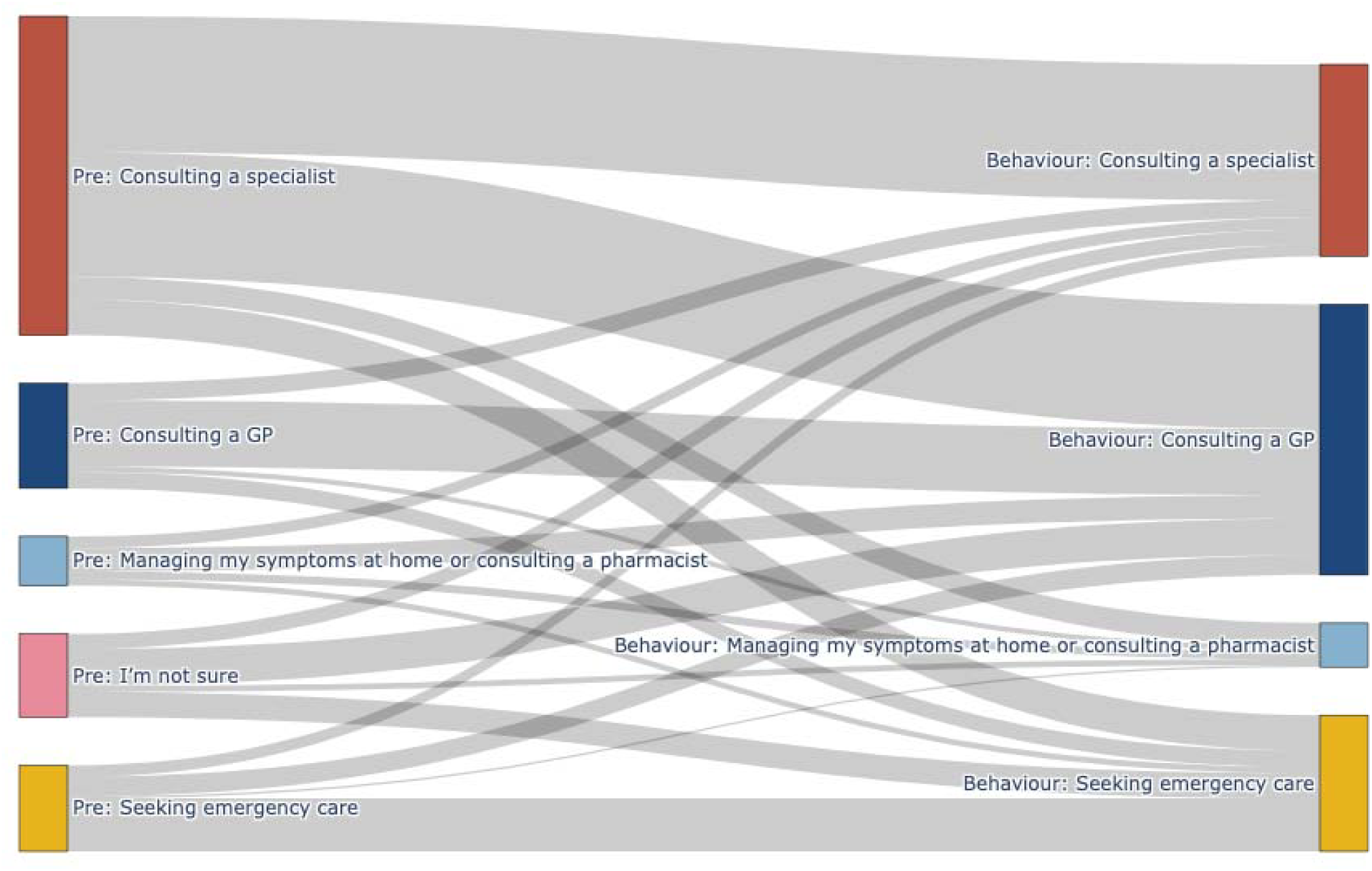
Sankey diagram depicting transitions from pre-assessment to actual behavior. Color scheme and node/link conventions are the same as in Figure 2A.

**Figure 2C:**
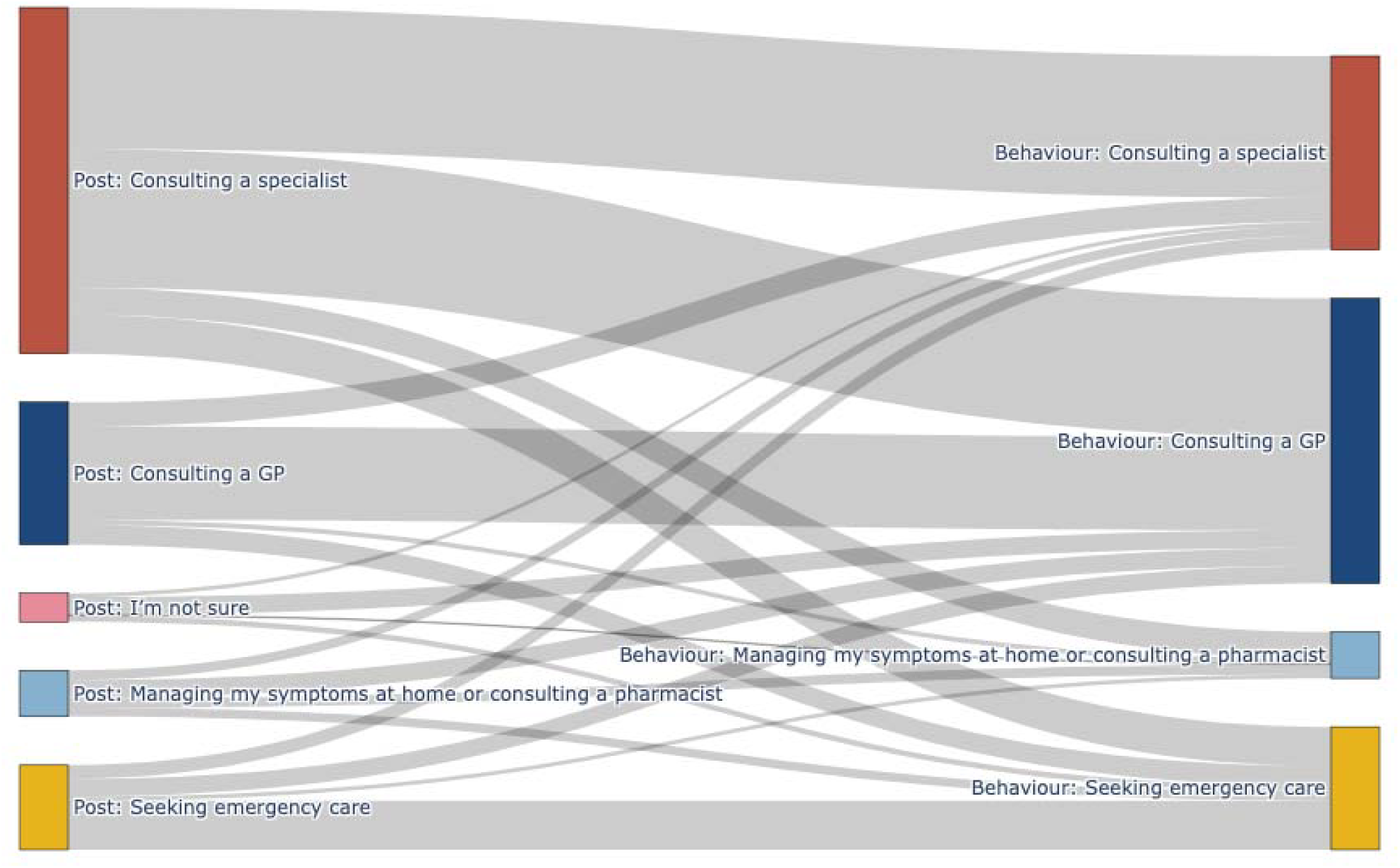
Sankey diagram depicting transitions from post-assessment to actual behavior. Color scheme and node/link conventions are the same as in Figure 2A and 2B.

### Participants Behavior

The proportion consulting a GP increased from 18.8□% to 41.8□% (p < 0.001). In contrast, specialist consultations declined from 57.1□% to 31.6□% (p < 0.001). Intentions to self-manage remained relatively stable (8.8□% vs. 6.7□%, p□= □0.18), while emergency care use rose from 15.4□% to 19.9□% (p□= □0.007) (Table S1).

The extent to which participants followed their initial care intentions varied notably across care pathways (Table S2). Only 18.4□% (9/55) of those who ultimately engaged in self-management had initially intended to do so, while 81.6□% (46/55) shifted into this pathway from a different intended level of care (Table S3).

Pre-assessment intention was the only statistically significant factor driving whether users stick with or revise their care plan (p < 0.001)(Table S5). The level of the cAI’s advice was associated with behaviour (χ^2^ = 25.44, df = 4, p = <0.001).

### Utilisation Impact

Among 717 participants for whom both pre-assessment intentions and observed behaviour were available, only 41% (294/717) followed through with the care level they had initially planned. 17.2□% (123/717) escalated to a higher-urgency option, while 28.9□% (207/716) de-escalated to a lower-urgency one, indicating a net shift toward lower-acuity care.

The overall distribution of care level shifted significantly from pre-assessment intentions to observed behaviour (p < 0.001), driven by a marked reduction in specialist care and increased GP consultations. The mean care intensity score decreased from 1.94 to 1.84 (p = 0.098). Users adhering to their initial plan selected higher-intensity care (mean behaviour score 2.09) than those who changed their plan (mean 1.63, p < 0.001).

### Appropriateness of plans for seeking care and observed behaviour

After completing the symptom assessment, the proportion of appropriate care intentions increased from 29.8□% (114/382) to 35.6□%(130/365) (p□= □0.076) (Table 4). 64.2□% (247/385) of behaviours were appropriate (p□< □0.001). Among initially unsure participants, 69□%(29/42) sought appropriate care. 77.0□% (296/385) of cases received the most appropriate urgency advice.

**Table 4.** Appropriateness of care plans before and after symptom assessment and of actual behaviour.

| Care-plan rating | Pre (n, %) | Post (n, %) | Actual Behavior (n, %) |
| --- | --- | --- | --- |
| **Appropriate** | 114 (29.8 %) | 130 (35.6 %) | 247 (64.2 %) |
| **Inappropriate** | 226 (59.2 %) | 216 (59.2 %) | 138 (35.8 %) |
| **Unsure** | 42 (11.0 %) | 19 (5.2 %) | — |
| **Total** | 382 | 365 | 385 |

### Emergency Department Utilisation and Appropriateness

Among users who initially planned to seek emergency care, 27.8□% (35/126) revised their intention immediately after the assessment, and 38.5□% (37/96) ultimately chose a different care level. Of these 96, 29 had a follow-up consultation within CUF with an appropriateness rating. In 27 cases (93.1□%), emergency care was not considered necessary, suggesting their decision to avoid the ED was clinically appropriate.

In both cases, the cAI recommended emergency care—one for pneumonia, the other for a suspected wrist fracture. Despite appropriate intentions and cAI advice, one participant saw a GP and the other a specialist.

Overall, 72.0□% of ED visits (95/132) were classified as appropriate. This included 55 visits judged clinically appropriate, and an additional 40 that involved ED-specific interventions such as in-hospital diagnostics, treatment, or admission.

Among appropriate ED visits, 56.8□% (54/95) were from patients who had not initially intended to seek emergency care. These visits involved slightly older patients (41.1□± □13.6 years) than inappropriate ones (36.0□± □12.9 years; p□= □0.053). When the cAI recommended emergency care, 82.1□% (32/39) of those visits were later rated as appropriate (Table S7). Based on the Manchester Triage Scale, 33.7□% of appropriate visits were classified as Urgent, compared to 8.1□% of inappropriate visits (p□= □0.003). Weekday visits were more often appropriate (74.7□% vs. 57.5□%; p□= □0.019).

### Generalisability

The largest difference between the intentions of the study population and the broader set of cAI users at CUF was in self-care intentions: post-assessment, 16.9□% of users in the overall population intended to self-manage, compared to just 7.3□% in the study population (p = 0.0066). No significant differences were observed in other care categories. The sex distribution however differed, with more male users in the study sample (41.8□%) than in the overall population (35.0□%)(Table S8).

Comparison of participants with and without follow-up data showed no major selection effects (Table S9). Users with follow-up were slightly older and more likely to have received high-acuity advice. They also showed a somewhat greater increase in planned GP consultations after the assessment, while changes in other care categories did not differ significantly.

## Discussion

The ESSENCE study evaluated a clinical AI integrated into Portugal’s largest private healthcare network to support appropriate patient healthcare utilisation. Following the cAI-based symptom assessment, patient uncertainty about managing their symptoms dropped significantly from 13% to 5%, and one in three patients immediately revised their intended care pathways. Behaviour assessed via EHR and follow-up surveys showed that ultimately 59% of participants altered their original care intentions: 17% increased urgency, 29% decreased urgency, and 13% became certain on how to manage their symptoms. Overall appropriateness of healthcare-seeking improved significantly from 30% pre-assessment to 64% based on actual behaviour, with ED utilisation being appropriate in 72% of cases. Among patients initially intending ED visits, 39% opted for alternative care options post-assessment, and follow-up data confirmed that emergency care was indeed unnecessary in 93% of these instances.

### The Role of AI in Patient Empowerment and Healthcare Utilisation

AI is increasingly recognised as a tool to support patients in navigating care decisions. By synthesising symptom data and offering personalised guidance, AI-driven tools can improve healthcare efficiency and appropriateness as seen in this study. Early evaluations focused on diagnostic accuracy, but interest is shifting toward real-world impacts on behaviour and outcomes. AI has influenced health behaviours in areas like hypertension, smoking cessation, and digital CBT for pain and anxiety^9–11^. However, evidence on how DFD tools affect real care decisions remains limited.

A study evaluating the integration of AI-supported virtual triage with live nurse triage in a Portuguese health insurance plan found 22.8% of patient-members revised their care intent after triage - compared to over 30% in the present study - but behaviour was only measured via outpatient bookings, excluding those choosing ED or self-care, with the focus of the paper being mainly on intent comparison^7^. Australia’s national virtual front door redirected 55% of ED-intent users to lower-acuity options, but adherence and appropriateness were not assessed^8^ (McMahon et al., 2025). A related LLM-based tool showed good technical performance but also lacked behavioural validation^12^.

Together, these studies reflect a growing interest in AI-supported navigation tools, but also highlight a common limitation: reliance on AI performance or stated intent rather than observed behaviour. In the present study, the poor agreement between post-assessment intent and actual behaviour (Cohen’s □ = 0.25) reinforces the need for robust, real-world evaluation to understand how DFD tools influence patient actions and system outcomes.

### Drivers of Behaviour Change

Understanding healthcare-seeking behaviour is inherently complex. In this study, one-third of users revised their care intentions immediately after reviewing the report, suggesting that the assessment process and structured information significantly influenced their initial decisions.

After this, users were shown mapped service options tailored to their condition and urgency, co-developed by CUF and the cAI provider. The high uptake of teleconsultations and same-day appointments suggests these actionable options strongly influenced behaviour. Prior research shows that immediacy and convenience often outweigh urgency in care decisions, and clear next steps likely acted as effective behavioural nudges^13–15^.

While triage recommendations guide decisions, patients also weigh whether advice fits their circumstances. Qualitative research shows people act on guidance that feels clear, reassuring, and feasible^4^. These patterns reflect behavioural economics models, where individuals often “satisfice” - choosing acceptable, if not optimal, options under uncertainty^16^. Patient engagement further improves when AI systems offer transparency, personalisation, and perceived empathy, creating a credible foundation for decision-making^14^.

### Appropriateness of Decision-Making

AI-driven DFDs aim to guide patients to appropriate care or safe self-management, improving outcomes and efficiency while reducing unnecessary or misdirected use. Yet, visit appropriateness is rarely evaluated - especially outside emergency care, where frameworks are more established.

These evaluations also are not straight forward. Retrospective judgments often lack insight into patient motivations or symptom progression. Tools focus on clinical effectiveness but often overlook barriers, reassurance needs, and cost considerations^17^. Even clinical ratings are subjective: Giannouchos et al. (2024) found 40–60% discordance in ED triage when comparing discharge diagnoses to presenting complaints^18^.

Frameworks also tend to assume a single “correct” option, ignoring that multiple care paths may be reasonable. In practice, ambiguity, overlapping symptoms, and context shape behaviour. In ESSENCE, weekend visits were more often rated inappropriate - reflecting how convenience and wait times can outweigh clinical logic, echoing Kraaijvanger et al. (2016)^19^.

Further insights come from experimental research outside clinical settings. In a randomised trial, Kopka et al. (2024) showed that laypeople using a cAI improved triage accuracy (53.2% to 64.5%), tripled correct self-care recognition (13.1% to 36.9%), and increased emergency detection by 29%, without raising undertriage^4^. Users were also more likely to revise urgency (27% vs. 17% with a general-purpose LLM), supporting ESSENCE’s behavioural findings.

While few studies assess AI-supported decision-making before care is sought, ED-based evaluations exist. Taylor et al. (2025) studied an AI triage tool across U.S. EDs, reporting better critical illness detection (78.8% to 83.1%) and faster care^20^. Rather than binary labels, the study used downstream outcomes - like critical care need and hospitalisation - as proxies for quality.

### Strengths of the Study

The ESSENCE study is among the first prospective real-world evaluations of cAI integrated into a healthcare system to influence patient utilisation. Its methodological strength lies in combining self-reported intentions with objective follow-up data from electronic health records and email surveys with a high response rate, allowing robust triangulation of planned versus actual behaviour - addressing a common limitation of digital health research. Conducted within Portugal’s largest private healthcare network, the study benefited from a large, heterogeneous user base spanning the full spectrum of care from self-management to specialist and emergency visits, and uniquely applied structured appropriateness criteria beyond emergency settings. The rigorous prospective design, aligned with ISO 14155:2020 good clinical practice guidelines, one-year data collection, and inclusion of diverse adult specialties enhance generalisability and provide a strong foundation for understanding clinical AI’s real-world impact on healthcare utilisation.

## Limitations

Several design aspects may introduce selection effects. Only users who consented to share their symptom report - after viewing bookable care pathways - were included, likely skewing the sample toward those already intent on seeking care. This may explain the lower rate of post-assessment self-care intentions (7.3%) compared to the broader cAI user base (16.9%).

Follow-up data were also uneven: while care at CUF facilities was fully captured via EHR, out-of-network care relied on voluntary email surveys, likely underreporting self-care and avoided visits - especially among those lost to follow-up. Additionally, appropriateness ratings were assessed using different methods by care level, and self-care decisions could not be evaluated. This hybrid approach supports exploratory comparison but limits definitive conclusions across all settings. Finally, the study took place in a well-resourced private system with faster access and an insurance- or self-pay model, which may limit generalisability to lower-resource or public healthcare contexts.

## Conclusion and Outlook

The ESSENCE study provides real-world evidence that clinical AI can support more confident and appropriate patient decision-making. By combining personalised symptom assessment with actionable service options, the integration reduced uncertainty, shifted care intentions, and promoted more appropriate use - highlighting AI’s potential as a behavioural nudge within care pathways.

Importantly, AI’s impact extends beyond output accuracy to how information is structured, perceived, and acted upon. This underscores the need for user-centred design and thoughtful integration into healthcare systems.

Future research should examine which report elements and service links drive behaviour, using methods like A/B testing and user interviews. External influences - such as symptom evolution and social input - must also be considered. Broader evaluations in public and underserved settings are needed to assess generalisability, alongside health economic studies quantifying cost and system impact.

## Data Availability

Data collected for the study, including individual participant data and a data dictionary defining each field in the set, will be made available to others after publication upon reasonable request, subject to approval. Requests for access should be made to the study team at Ada Health. After approval, a signed data sharing agreement will be required before data release.

## Contributors

FC, SG, PF and MSM were responsible for conceptualisation and methodology. PF, MP, FC, NK, TM, AP and MS contributed to investigation, project administration, supervision, validation, software and data curation. FC was additionally responsible for formal analysis and visualisation. MS was also responsible for resources and review and editing while FC additionally led writing the original draft in collaboration with all co-authors. AL contributed to project administration, supervision and writing - review and editing. All authors had full access to aggregated data in the study and contributed to the decision to submit the manuscript for publication.

## Acknowledgements

This work was supported by the Federal Ministry of Education and Research (Bundesministerium für Bildung und Forschung) through the European Union-financed NextGenerationEU program under grant number 16KISA100K, project PATH—“Personal Mastery of Health and Wellness Data.”

