## Supplementary Appendix for "From Advice to Action: Real-World Behaviour of Patients Using an Integrated Clinical AI for Navigating the Healthcare System"

ESSENCE healthcare utilisation publication - APPENDIX

### Table of Contents:

**Table S1.** Change in care intentions and behaviour — McNemar test results	1

**Table S2.** Concordance between pre-assessment intention and observed behaviour, by care pathway	2

**Table S3A and S3B.** Contingency table: Pre-assessment intention vs. Observed behavior	2

**Table S4A and S4B.** Contingency table: Post-assessment intention vs. Observed behavior	3

**Table S5.** Univariate Associations with Plan Change	4

**Table S6**. Contingency Tables with granular categories. A. Pre-Post assessment, B. Pre-Assessment vs. Behavior C. Post-assessment vs. Behavior	4

**Table S7.** Characteristics of emergency department (ED) visits by appropriateness classification	6

**Table S8.** Comparison of Study Population with Overall Consented Population (Excluding Skipped Responses) A. Change in Care Intentions Before and After Assessment, B. Distribution of cAI Advice Level (Merged Categories), C. Sex Distribution	7

**Table S9.** Change in Care Intentions (Pre → Post) for Participants With Known vs. Unknown Behaviour	8

**Table S10**. Unmerged care option distributions for Pre-assessment intention, Post-assessment intention, and Behavior	9

###### **Table S1.** Change in care intentions and behaviour — McNemar test results

| **A. Pre-assessment to Post-assessment shifts per category** | | | | | | | |
| --- | --- | --- | --- | --- | --- | --- | --- |
| **Care option** | **Pre n** | **Pre %** | **Post n** | **Post %** | **Pre→No (b)** | **No→Post (c)** | **McNemar p (two-sided)** |
| Consulting a specialist | 709 | 53.0 % | 767 | 57.3 % | 96 | 154 | <0.001 |
| Consulting a GP | 238 | 17.8 % | 263 | 19.7 % | 83 | 108 | 0.0822 |
| I’m not sure | 168 | 12.6 % | 67 | 5.0 % | 138 | 37 | <0.001 |
| Seeking emergency care | 126 | 9.4 % | 144 | 10.8 % | 35 | 53 | 0.0694 |
| Managing my symptoms at home / consulting a pharmacist | 97 | 7.2 % | 97 | 7.2 % | 59 | 59 | 1 |
| **B. Pre-assessment to Behavior shifts per category** | | | | | | | |
| **Care option** | **Pre n** | **Pre %** | **Behaviour n** | **Behaviour %** | **Pre→No (b)** | **No→Beh (c)** | **McNemar p (two-sided)** |
| Consulting a GP | 117 | 18.8 % | 261 | 41.8% | 43 | 187 | <0.001 |
| Consulting a specialist | 356 | 57.1 % | 197 | 31.6% | 204 | 45 | <0.001 |
| Managing my symptoms at home / consulting a pharmacist | 55 | 8.8 % | 42 | 6.7 % | 46 | 33 | 0.1766 |
| Seeking emergency care | 96 | 15.4 % | 124 | 19.9 % | 37 | 65 | 0.00721 |

###### **Table S2.** Concordance between pre-assessment intention and observed behaviour, by care pathway

| **Pre-assessment intention** | **Total (n)** | **Same (n)** | **% Same** |
| --- | --- | --- | --- |
| Consulting a GP | 117 | 74 | 63.20% |
| Consulting a specialist | 356 | 152 | 42.70% |
| Managing my symptoms at home / consulting a pharmacist | 55 | 9 | 16.40% |
| Seeking emergency care | 96 | 59 | 61.50% |

###### **Table S3A and S3B.** Contingency table: Pre-assessment intention vs. Observed behavior, n=717, 4 excluded as skipped pre-assessment question

| **Pre-assessment intention** | **Behavior** | | | | |
| --- | --- | --- | --- | --- | --- |
|  | **Self-care** | **Consulting a GP** | **Consulting a specialist** | **Seeking emergency care** | **Total** |
| **Self-care** | 9 (16.4%) | 26 (47.3%) | 13 (23.6%) | 7 (12.7%) | 55 |
| **GP** | 6 (5.1%) | 74 (63.2%) | 19 (16.2%) | 18 (15.4%) | 117 |
| **Specialist** | 25 (7.0%) | 139 (39.0%) | 152 (42.7%) | 40 (11.2%) | 356 |
| **ED** | 2 (2.1%) | 22 (22.9%) | 13 (13.5%) | 59 (61.5%) | 96 |
| **I’m not sure** | 7 (7.5%) | 41 (44.1%) | 17 (18.3%) | 28 (30.1%) | 93 |
| **Total** | 49 | 302 | 214 | 152 | 717 |

| **Pre-assessment intention** | **Behavior** | | | | |
| --- | --- | --- | --- | --- | --- |
|  | **Self-care** | **Consulting a GP** | **Consulting a specialist** | **Seeking emergency care** | **Total** |
| **Self-care** | 9 (18.4%) | 26 (8.6%) | 13 (6.1%) | 7 (4.6%) | 55 |
| **GP** | 6 (12.2%) | 74 (24.5%) | 19 (8.9%) | 18 (11.8%) | 117 |
| **Specialist** | 25 (51.0%) | 139 (46.0%) | 152 (71.0%) | 40 (26.3%) | 356 |
| **ED** | 2 (4.1%) | 22 (7.3%) | 13 (6.1%) | 59 (38.8%) | 96 |
| **I’m not sure** | 7 (14.3%) | 41 (13.6%) | 17 (7.9%) | 28 (18.4%) | 93 |
| **Total** | 49 (100%) | 302 (100%) | 214 (100%) | 152 (100%) | 717 |

###### **Table S4A and S4B.** Contingency table: Post-assessment intention vs. Observed behavior, n = 659

| **Post-assessment intention** | **Behavior** | | | | |
| --- | --- | --- | --- | --- | --- |
|  | **Self-care** | **Consulting a GP** | **Consulting a specialist** | **Seeking emergency care** | **Total** |
| **Self-care** | 10 (21.3%) | 18 (38.3%) | 10 (21.3%) | 9 (19.1%) | 47 |
| **Consulting a GP** | 5 (3.4%) | 95 (65.5%) | 25 (17.2%) | 20 (13.8%) | 145 |
| **Consulting a specialist** | 27 (7.7%) | 141 (40.1%) | 144 (40.9%) | 40 (11.4%) | 352 |
| **Seeking emergency care** | 4 (4.7%) | 18 (20.9%) | 14 (16.3%) | 50 (58.1%) | 86 |
| **I’m not sure** | 1 (3.4%) | 18 (62.1%) | 4 (13.8%) | 6 (20.7%) | 29 |
| **Total** | 47 (100%) | 290 (100%) | 197 (100%) | 125 (100%) | 659 |

| **Post-assessment intention** | **Behavior** | | | | |
| --- | --- | --- | --- | --- | --- |
|  | **Self-care** | **Consulting a GP** | **Consulting a specialist** | **Seeking emergency care** | **Total** |
| **Self-care** | 10 (21.3%) | 18 (6.2%) | 10 (5.1%) | 9 (7.2%) | 47 |
| **Consulting a GP** | 5 (10.6%) | 95 (32.8%) | 25 (12.7%) | 20 (16.0%) | 145 |
| **Consulting a specialist** | 27 (57.4%) | 141 (48.6%) | 144 (73.1%) | 40 (32.0%) | 352 |
| **Seeking emergency care** | 4 (8.5%) | 18 (6.2%) | 14 (7.1%) | 50 (40.0%) | 86 |
| **I’m not sure** | 1 (2.1%) | 18 (6.2%) | 4 (2.0%) | 6 (4.8%) | 29 |
| **Total** | 47 (100%) | 290 (100%) | 197 (100%) | 125 (100%) | 659 |

###### **Table S5**. Univariate Associations with Plan Change (Same vs Changed, N = 623)

| **Predictor** | **Levels (% changed)** | **Test** | **Statistic** | **p-value** |
| --- | --- | --- | --- | --- |
| **Age** | Mean same = 40.31 y; Mean changed = 40.00 y | Mann–Whitney U | U = 51922.00 | 0.112 |
| **Number of presenting symptoms entered** | Mean same = 2.42; Mean changed = 2.22 | Mann–Whitney U | U = 49751.00 | p = 0.516 |
| **Sex** | Female 54.7 % vs Male 50.2 % | χ² test | χ² = 1.04 | p = 0.307 |
| **Case Advice merged** | Self-care 56.4 %, Primary-care 52.3 %, Emergency 52.1 % | χ² test | χ²₂ = 0.47 | 0.792 |
| **Pre-assessment intention merged** | Self-care 83.6 %, GP 38.8 %, Specialist 57.3 %, Emergency 38.5 % | χ² test | χ² = 43.81 | **<0.001** |

###### **Table S6.** Contingency Tables with granular categories. **A.** Pre-Post assessment, **B.** Pre-Assessment vs. Behavior **C**. Post-assessment vs. Behavior

| **Pre-assessment intention** | **Post-assessment intention** | | | | | | | |
| --- | --- | --- | --- | --- | --- | --- | --- | --- |
|  | **Consulting a GP in person** | **Consulting a GP online or by phone** | **Consulting a specialist in person** | **Consulting a specialist online or by phone** | **I’m not sure** | **Managing my symptoms at home or consulting a pharmacist** | **Seeking emergency care** | **Skip** |
| **Consulting a GP in person** | 46 | 4 | 23 | 1 | 5 | 7 | 4 | 2 |
| **Consulting a GP online or by phone** | 6 | 99 | 10 | 14 | 5 | 8 | 6 | 5 |
| **Consulting a specialist in person** | 24 | 2 | 415 | 19 | 4 | 12 | 16 | 37 |
| **Consulting a specialist online or by phone** | 3 | 17 | 24 | 156 | 5 | 7 | 6 | 22 |
| **I’m not sure** | 12 | 21 | 37 | 27 | 30 | 23 | 18 | 21 |
| **Managing my symptoms at home or consulting a pharmacist** | 7 | 15 | 10 | 11 | 13 | 38 | 3 | 10 |
| **Seeking emergency care** | 2 | 5 | 11 | 9 | 5 | 2 | 91 | 19 |
| **Skip** | 0 | 1 | 9 | 0 | 1 | 2 | 0 | 3 |

Contingency Table Pre-assessment & Behavior (all values)

| **Pre-assessment intention** | **Behavior** | | | | | | | |
| --- | --- | --- | --- | --- | --- | --- | --- | --- |
|  | **Consulting a GP in person** | **Consulting a GP online or by phone** | **Consulting a specialist in person** | **Consulting a specialist online or by phone** | **EHR file not accessible** | **Managing my symptoms at home or consulting a pharmacist** | **No consultation at CUF** | **Seeking emergency care** |
| **Consulting a GP in person** | 16 | 8 | 11 | 0 | 1 | 2 | 47 | 7 |
| **Consulting a GP online or by phone** | 8 | 42 | 8 | 0 | 1 | 4 | 79 | 11 |
| **Consulting a specialist in person** | 35 | 27 | 118 | 3 | 19 | 18 | 278 | 30 |
| **Consulting a specialist online or by phone** | 15 | 62 | 23 | 8 | 6 | 7 | 109 | 10 |
| **I’m not sure** | 19 | 22 | 17 | 0 | 3 | 7 | 93 | 28 |
| **Managing my symptoms at home or consulting a pharmacist** | 5 | 21 | 11 | 2 | 3 | 9 | 49 | 7 |
| **Seeking emergency care** | 8 | 14 | 13 | 0 | 1 | 2 | 48 | 59 |
| **Skip** | 0 | 2 | 1 | 0 | 0 | 1 | 12 | 0 |

Contingency Table Post-assessment & Behavior (all values)

| **Post-assessment intention** | **Behavior** | | | | | | | |
| --- | --- | --- | --- | --- | --- | --- | --- | --- |
|  | **Consulting a GP in person** | **Consulting a GP online or by phone** | **Consulting a specialist in person** | **Consulting a specialist online or by phone** | **EHR file not accessible** | **Managing my symptoms at home or consulting a pharmacist** | **No consultation at CUF** | **Seeking emergency care** |
| **Consulting a GP in person** | 23 | 12 | 11 | 0 | 3 | 2 | 39 | 10 |
| **Consulting a GP online or by phone** | 13 | 47 | 14 | 0 | 3 | 3 | 74 | 10 |
| **Consulting a specialist in person** | 33 | 29 | 117 | 2 | 15 | 21 | 292 | 29 |
| **Consulting a specialist online or by phone** | 16 | 63 | 19 | 6 | 6 | 6 | 111 | 11 |
| **I’m not sure** | 5 | 13 | 4 | 0 | 0 | 1 | 39 | 6 |
| **Managing my symptoms at home or consulting a pharmacist** | 4 | 14 | 8 | 2 | 2 | 10 | 50 | 9 |
| **Seeking emergency care** | 9 | 9 | 14 | 0 | 2 | 4 | 56 | 50 |
| **Skip** | 3 | 11 | 15 | 3 | 3 | 3 | 54 | 27 |

###### **Table S7.** Characteristics of emergency department (ED) visits by appropriateness classification (N = 132).

| **Characteristic** | **Inappropriate (N = 37)** | **Appropriate (Y = 95)** | **p-value** |
| --- | --- | --- | --- |
| **Age (years)** | 36.0 ± 12.9 (n = 37) | 41.1 ± 13.6 (n = 95) | 0.053ⁿ |
| **Sex** |  |  | 0.935ᶜ |
| Female | 22 (59.5 %) | 59 (62.1 %) |  |
| Male | 15 (40.5 %) | 36 (37.9 %) |  |
| **Pre-assessment intention** |  |  | 0.743ᶜ |
| Consulting a GP | 4 (10.8 %) | 9 ( 9.5 %) |  |
| Consulting a specialist | 10 (27.0 %) | 26 (27.4 %) |  |
| I’m not sure | 9 (24.3 %) | 15 (15.8 %) |  |
| Self-care / pharmacy | 2 ( 5.4 %) | 4 ( 4.2 %) |  |
| Seeking emergency care | 12 (32.4 %) | 41 (43.2 %) |  |
| **Manchester Triage Scale** |  |  | **0.003ᶠ** |
| Less or Non-Urgent | 30 (81.1 %) | 58 (61.1 %) |  |
| Urgent & Very Urgent | 3 ( 8.1 %) | 32 (33.7 %) |  |
| **Presentation day** |  |  | **0.019ᶠ** |
| Weekday | 19 (51.4 %) | 71 (74.7 %) |  |
| Other (Weekend/Holiday) | 17 (45.9 %) | 23 (24.2 %) |  |
| **cAI emergency recommendation** |  |  | 0.136ᶠ |
| Not emergency | 30 (81.1 %) | 63 (66.3 %) |  |
| Emergency recommended | 7 (18.9 %) | 32 (33.7 %) |  |
| ⁿ Student’s t-test ᶜ Chi-square test ᶠ Fisher’s exact test. |  |  |  |

###### **Table S8.** Comparison of Study Population with Overall Consented Population (Excluding Skipped Responses) **A.** Change in Care Intentions Before and After Assessment, **B.** Distribution of cAI Advice Level (Merged Categories), **C.** Sex Distribution

| **Intentions** | **Study Pre %** | **Study Post %** | **Study Δ pp** | **Overall Pre %** | **Overall Post %** | **Overall Δ pp** | **z-stat** | **p-value** |
| --- | --- | --- | --- | --- | --- | --- | --- | --- |
| Consulting a specialist (merged) | 52.9 % | 57.4 % | **4.55** | 41.7 % | 48.9 % | **7.15** | –1.26 | 0.206 |
| Consulting a GP (merged) | 16.9 % | 19.5 % | **2.69** | 15.9 % | 15.9 % | **0.02** | 1.69 | 0.091 |
| **I’m not sure** | 13.0 % | 5.0 % | **– 7.97** | 20.7 % | 9.7 % | **– 10.94** | 2.5 | 0.012 |
| Managing at home / pharmacist | 7.4 % | 7.3 % | **– 0.03** | 13.7 % | 16.9 % | **3.12** | –2.71 | 0.0066 |
| Seeking emergency care | 9.9 % | 10.7 % | **0.76** | 8.0 % | 8.6 % | **0.65** | 0.08 | 0.936 |

| **Category** | **Study %** | **Overall %** | **Δ (Study – Overall)** | **z-stat** | **p-value** |
| --- | --- | --- | --- | --- | --- |
| **Advice-level (n = 1 470 vs. 16 158)** | | | | | |
| Self-Care (combined) | 12.1 % | 14.9 % | – 2.8 pp | –3.08 | **0.002** |
| Primary Care (all tiers) | 74.8 % | 68.5 % | 6.3 pp | 5.31 | **1.1 × 10⁻⁷** |
| Emergency + Ambulance | 12.7 % | 16.7 % | – 4.0 pp | –4.41 | **1.1 × 10⁻⁵** |
| **Sex (n = 1 463 vs. 11 067)** | | | | | |
| Female | 57.7 % | 65.0 % | – 7.1 pp | –5.16 | **2.5 × 10⁻⁷** |
| Male | 41.8 % | 35.0 % | 7.1 pp | 5.16 | **2.5 × 10⁻⁷** |

###### **Table S9.** Change in Care Intentions (Pre → Post) for Participants With Known vs. Unknown Behaviour. Skips excluded from denominators; Δ pp = Post % – Pre %

| **Intention Category** | **Known Pre %** | **Known Post %** | **Δ pp<br>(Known)** | **Unknown Pre %** | **Unknown Post %** | **Δ pp<br>(Unknown)** | **z-stat** | **p-value** |
| --- | --- | --- | --- | --- | --- | --- | --- | --- |
| Consulting a specialist | 49.6 | 53.3 | **3.7** | 56 | 61.3 | **5.3** | –0.47 | 0.64 |
| Consulting a GP | 16.3 | 22 | **5.7** | 17.3 | 17.2 | **–0.1** | 2.01 | **0.044** |
| Managing at home / pharmacist | 7.7 | 7.1 | **–0.6** | 7 | 7.5 | **0.5** | –0.51 | 0.61 |
| Seeking emergency care | 13.4 | 13.1 | **–0.3** | 6.6 | 8.4 | **1.8** | –0.84 | 0.4 |
| I’m not sure | 13 | 4.4 | **–8.6** | 13 | 5.6 | **–7.4** | –0.57 | 0.57 |

| **Age (mean ± SD), Welch t : t = 2.78, p = 0.0057** | | | | |
| --- | --- | --- | --- | --- |
| Known : 39.5 ± 12.3 |  |  |  |  |
| Unknown : 37.7 ± 12.6 |  |  |  |  |
| **Sex: χ² = 2.12, p = 0.145** | | | | |
|  | Unknown Behavior | % | Known Behavior | % |
| Female | 417 | 56.0 | 431 | 60.0 |
| Male | 327 | 44.0 | **288** | 40.0 |
| Total | **744** |  | **719** |  |
| **Advice level (merged): χ² = 8.44, p = 0.0147** | | | | |
|  | Unknown Behavior | % | Known Behavior | % |
| Self Care | 83 | 11.2 | 95 | 13.2 |
| Primary Care | 582 | 78.2 | 517 | 71.9 |
| Emergency Care | 79 | 10.6 | 107 | 14.9 |
| Total | **744** |  | **719** |  |

###### **Table S10.** Unmerged care option distributions for Pre-assessment intention, Post-assessment intention, and Behavior (n, %)

| **Care Level** | **Pre (n, %)** | **Post (n, %)** | **Behavior (n, %)** |
| --- | --- | --- | --- |
| Consulting a specialist in person | 528 (35.9%) | 538 (36.6%) | 202 (13.7%) |
| Consulting a specialist online or by phone | 240 (16.3%) | 238 (16.2%) | 13 (0.9%) |
| I’m not sure | 189 (12.9%) | 68 (4.6%) | 0 (0.0%) |
| Consulting a GP online or by phone | 153 (10.4%) | 164 (11.2%) | 198 (13.5%) |
| Seeking emergency care | 145 (9.9%) | 144 (9.8%) | 152 (10.3%) |
| Managing my symptoms at home or consulting a pharmacist | 107 (7.3%) | 99 (6.7%) | 50 (3.4%) |
| Consulting a GP in person | 92 (6.3%) | 100 (6.8%) | 106 (7.2%) |
| Skip | 16 (1.1%) | 119 (8.1%) | 0 (0.0%) |
| No consultation at CUF | — | — | 715 (48.6%) |
| EHR file not accessible | — | — | 34 (2.3%) |
